# A compartmental Mathematical model of COVID-19 intervention scenarios for Mumbai

**DOI:** 10.1101/2022.02.28.22271624

**Authors:** Avaneesh Singh, Manish Kumar Bajpai

## Abstract

A new mathematical method with an outstanding potential to predict the incidence of COVID-19 diseases has been proposed. The model proposed is an improvement to the SEIR model. In order to improve the basic understanding of disease spread and outcomes, four compartments included presymptomatic, asymptomatic, quarantine hospitalized and hospitalized. We have studied COVID-19 cases in the city of Mumbai. We first gather clinical details and fit it on death cases using the Lavenberg-Marquardt model to approximate the various parameters. The model uses logistic regression to calculate the basic reproduction number over time and the case fatality rate based on the age-category scenario of the city of Mumbai. Two types of case fatality rate are calculated by the model: one is CFR daily, and the other is total CFR. The total case fatality rate is 4.2, which is almost the same as the actual scenario. The proposed model predicts the approximate time when the disease is at its worst and the approximate time when death cases barely arise and determines how many hospital beds in the peak days of infection would be expected. The proposed model outperforms the classic ARX, SARIMAX and the ARIMA model. And It also outperforms the deep learning models LSTM and Seq2Seq model. To validate results, RMSE, MAPE and R squared matrices are used and are represented using Taylor diagrams graphically.

## 1 Introduction

The whole world is currently facing a novel coronavirus (COVID-19) pandemic that began in December 2019 in Wuhan City of China as an outbreak of pneumonia of unknown cause [1–3]. Many places have been greatly affected by this disease, and Mumbai is one of them. Mumbai is India’s financial capital and the capital of the state of Maharashtra. One of the epicenters of COVID-19 in India is Mumbai. As of 2019, Mumbai is the second-most populous city in the country after Delhi, according to the United Nations, and the seventh-most populous city in the world with a population of about 20 million[23]. As per the 2011 Indian Government population census, Mumbai was India’s most populous city with an estimated citywide population of 12.5 million living under the Greater Mumbai Municipal Corporation [24]. The model has been parameterized using available mortality data from COVID-19, which is more accurate than case data. In Mumbai, as of 30 November 2020, 280,818 confirmed cases and 11559 confirmed deaths due to COVID-19 had been reported.

It is an important decision for India to come up with a non-pharmaceutical control strategy such as a 40-day national lockdown to extend the higher phases of COVID-19 and to prevent severe loads on its public health system. On 11 March 2020, the pandemic novel coronavirus 2019 had been first confirmed in Mumbai. The Indian government has introduced a preventive social distance measure in order to avoid large-scale population movements that can accelerate the spread of the disease. On 22 March 2020, India’s government introduced a voluntary 14-hour public curfew. In addition, on midnight 24 March 2020, India’s prime minister also ordered a national lockdown to slow COVID-19 spread.

Due to absence of specific vaccine, strategies for controlling and mitigating the burden of the pandemic are focused on non-pharmaceutical interventions, such as social-distancing, contact-tracing, quarantine, isolation, and the use of facemasks in public, tracing, quarantine hospitalized of asymptomatic cases, and hospitalization of confirmed cases etc. are essential to control the spread of COVID-19 [4].

We are proposing a mathematical model that predicts Mumbai’s COVID-19 dynamics. The dynamics of diseases transmission can be achieved by mathematical modelling using differential equation system [5]. For the COVID-19 outbreak, several mathematical modelling studies have been done [6–13, 45, 46]. In this study, with epidemic data up to 30 October 2020, we propose a compartmental mathematical model to predict and control the transmission dynamics of the COVID-19 pandemic for Mumbai. We compute the basic reproduction number R_0_, which is used to study the predictions and simulations of the model. When epidemiological disease dynamics of transmission are not known, mathematical models play an important role in estimating the worst and best case scenarios. The main objective of the preventive techniques is to preserve the basic reproduction number R_0_ below one, in order to control the further development of infection, while the main objective of the mitigation policy is to determine the effect of the outbreak [10]. In order to understand the transmission dynamics of peculiar epidemiological traits of COVID-19, some mathematical models have recently been investigated, and some of these are listed in our references [13–22]. The modelling of COVID-19 pandemics is no new and in the absence of any effective therapeutics and licenced vaccine, most authors have focused on non-pharmaceutical interventions. Within that context, A mathematical model to study the transmission dynamics of the COVID-19 pandemic was studied by A. Atangana[13], using a system of fractional differential equations using the lockdown effect. In order to obtain epidemiological status from patients, Tang and colleagues [14] proposed the COVID-19 model, which is very high for infectious diseases, and calculated the basic reproduction number 6.47. The new SIDARTHE model for the COVID-19 pandemic has been developed by Giordano and colleagues[16], predicting that restrictive social separation can reduce the prevalence of coronavirus among humans. In order to investigate transmitting dynamics of the COVID-19, the Sarkar and Khajanchi [15,18] proposed and analyzed a mathematical model with true data, respectively, from India and certain states of India. The authors performed the short-term and long-term prediction based on estimated model parameters. In an extensive version of the classical SEIR model to study infectious diseases intervention strategies that incorporates the fact that asymptomatic and pre-symptomatic persons infected are believed to be a major role in the dynamics of COVID-19 outbreak transmission [17].

Fig. 1 shows the distribution of COVID-19 disease in the Maharashtra state. Case distribution is more in Mumbai, Thane, Nagpur, Nashik and Pune city.

**Figure 1:**
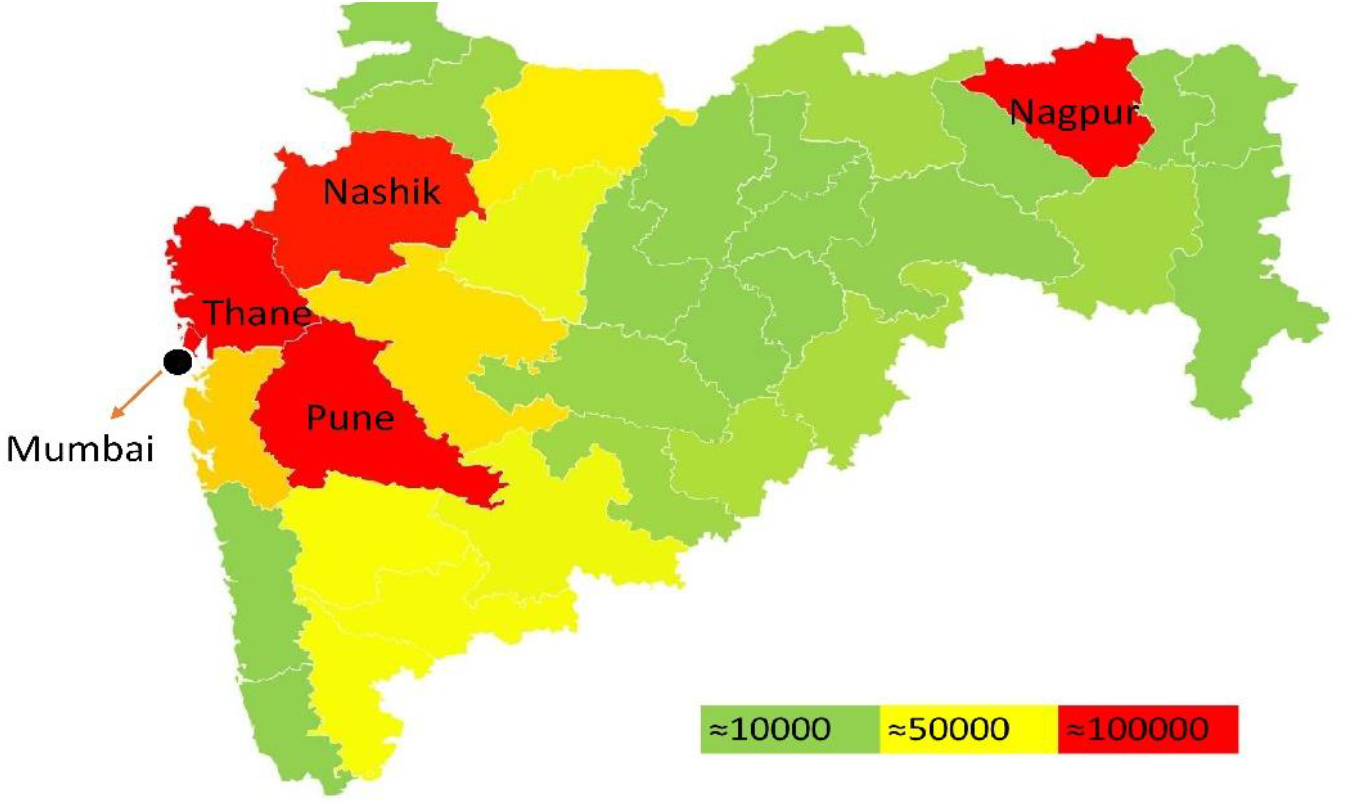
Geographically COVID-19 case distribution in the state of Maharashtra

If we compare Mumbai City with the United States of America, then population wise Mumbai city is nearly same as the New York state. Fig. 2 shows the population wise compare of Indian places with the USA states.

**Figure 2:**
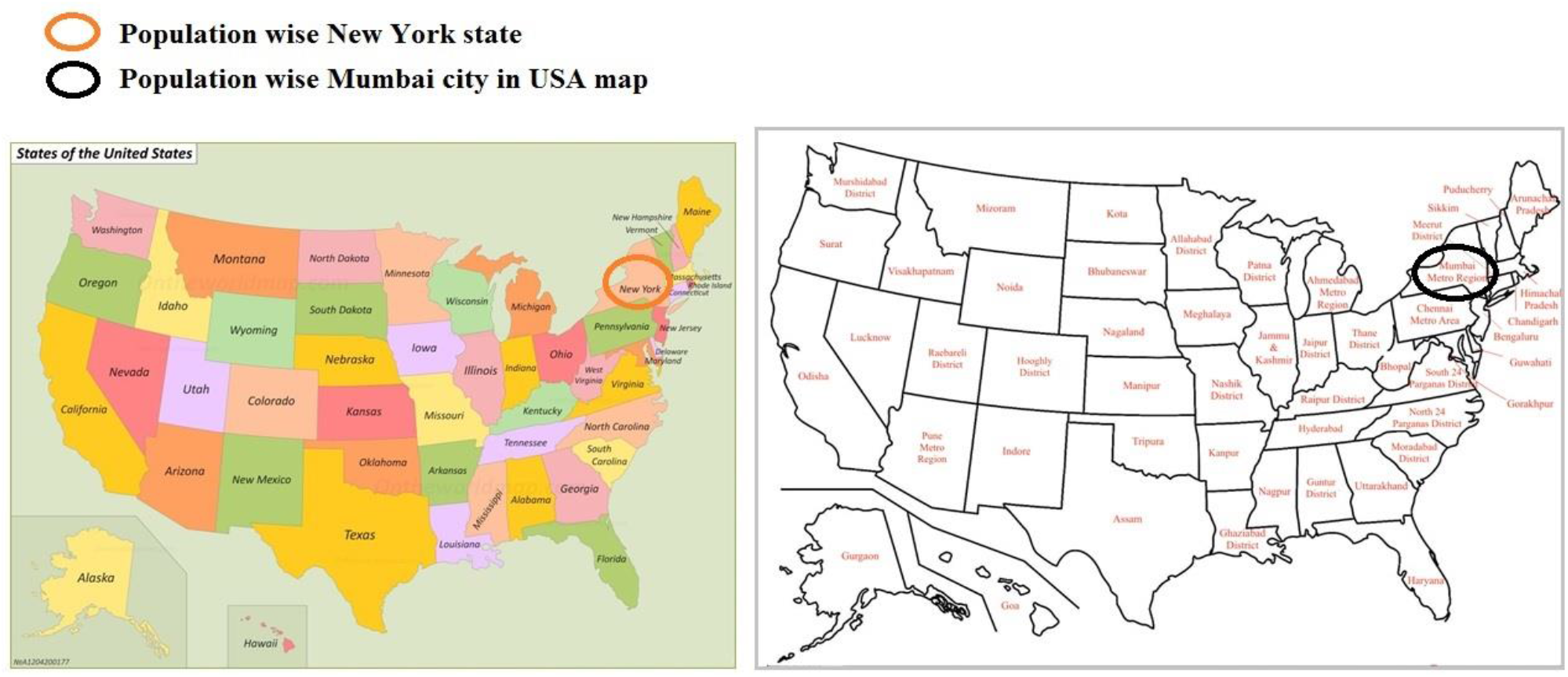
Population wise comparison of Indian places with USA sates Figure 2 reference link: https://ontheworldmap.com/usa/usa-states-map.html

The 2019 pandemic novel coronavirus was first confirmed in Mumbai on March 11, 2020. Moreover, Table 1 includes the important events happened in Mumbai city related to COVID-19.

**Table 1:** Important events in Mumbai related COVID-19

| S. No. | Date | Mumbai important Events |
| --- | --- | --- |
| 1 | 11-03-2020 | First Positive test result |
| 2 | 13-03-2020 | The Government of Maharashtra declared the outbreak an epidemic in the city of Mumbai |
| 3 | 14-03-2020 | Ban on public gatherings and events |
| 4 | 16-03-2020 | A three-year - old child and her mother were tested positive |
| 5 | 17-03-2020 | First death case because of COVID-19 |
| 6 | 20-03-2020 | Closure of all workplaces barring essential services |
| 7 | 22-03-2020 | Imposition of Section 144 and lockdown |
| 8 | 23-03-2020 | Curfew and border seal-off in Mumbai |
| 9 | 25-03-2020 | Nationwide lockdown until 14 April |
| 10 | 26-03-2020 | First Woman death due to COVID-19 |
| 11 | 11-04-2020 | Lockdown extended until 30 April |
| 12 | 14-04-2020 | Nationwide lockdown until 3 May |
| 13 | 20-04-2020 | 53 journalist found positive |
| 14 | 21-04-2020 | Cases surpassed 3000 |
| 15 | 01-05-2020 | Nationwide lockdown until 17 May |
| 16 | 07-05-2020 | Total cases in Mumbai crosses 10,000 mark |
| 17 | 16-05-2020 | 884 cases were reported in the Mumbai city |

The organization of this manuscript is as follows: Section 1 introduces COVID-19 and explains the importance of this research. The mathematical model and its schematic representation have been proposed in Section 2. The qualitative characteristics of the model and data analysis have been discussed in Section 3. Numerical simulations and results based on the estimated values of the parameters and the discussion are discussed in Section 4. Finally, the conclusion and some future work have been discussed in Section 5.

## 2. Proposed methodology

How the virus spreads from one location to another is called virus transmission. The transmission of infection occurs when an infectious person or source of infection infects susceptible people. There have to be several events that allow viruses to cause infections in susceptible people. This is called the Chain of Infection.

Two types of transmission mode are:

1. Direct Transmission
2. Indirect Transmission

### Direct Transmission

In direct transmission, an infectious person, through direct contact or droplet spread, is transmitted from the source of infection to susceptible people.

### Indirect Transmission

In Indirect transmission, an infectious person, through indirect contact or Airborne or Vehicle-borne, is transmitted from the source of infection to susceptible people.

Fig. 3 shows the chain of infection from an infected person to a susceptible person.

**Figure 3:**
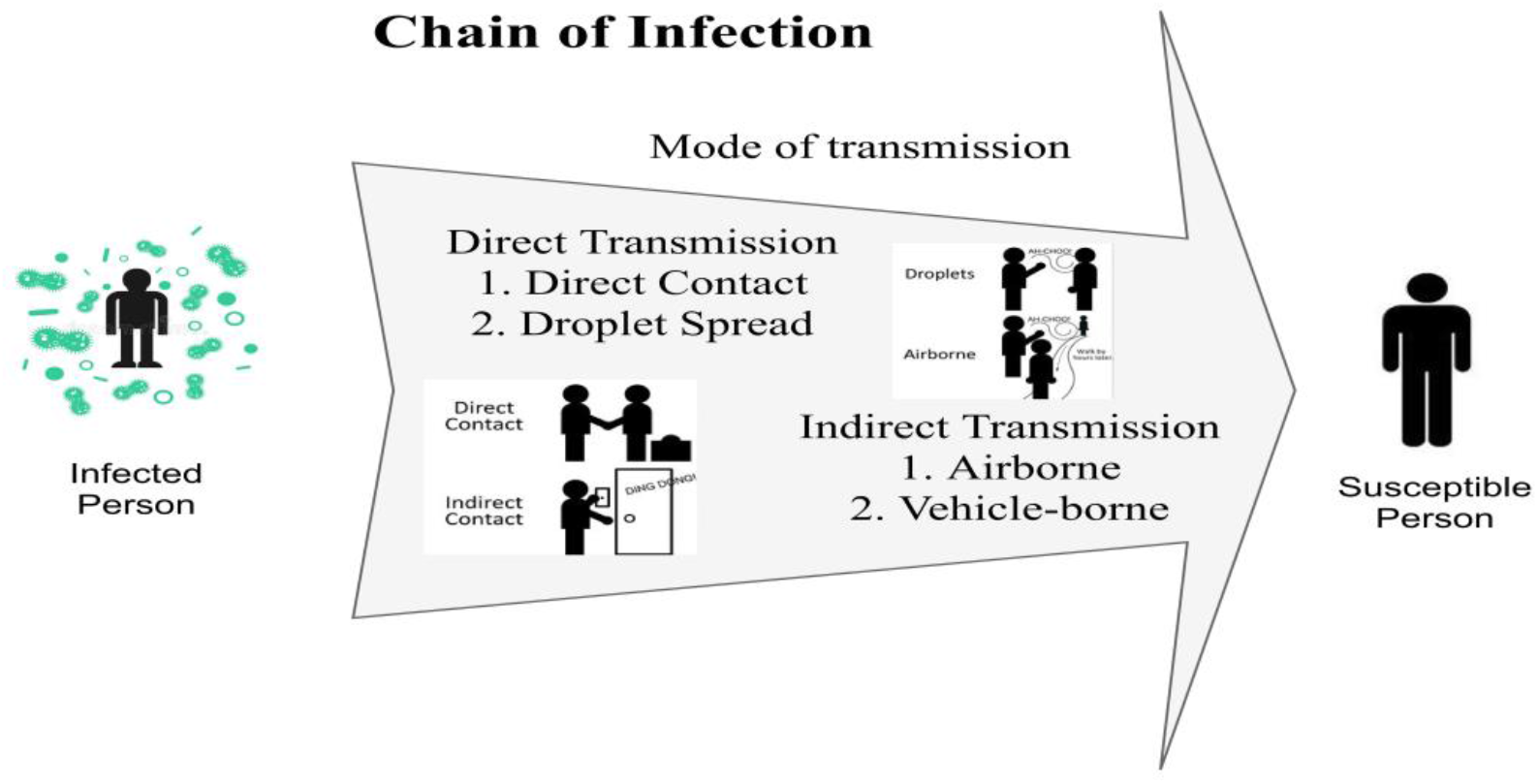
Chain of Infection

Infection transmission from one infected person to susceptible persons is shown in Fig. 4 The individual may be infected either symptomatically, asymptomatically, or pre-symptomatically. The lower box suggests that the majority of cases are either moderate or asymptomatic. Severe cases are relatively lesser than mild and asymptomatic cases but more than critical ones. Some cases are initially moderate, but with time, severe or critical; they need hospital beds or ICU beds.

**Figure 4:**
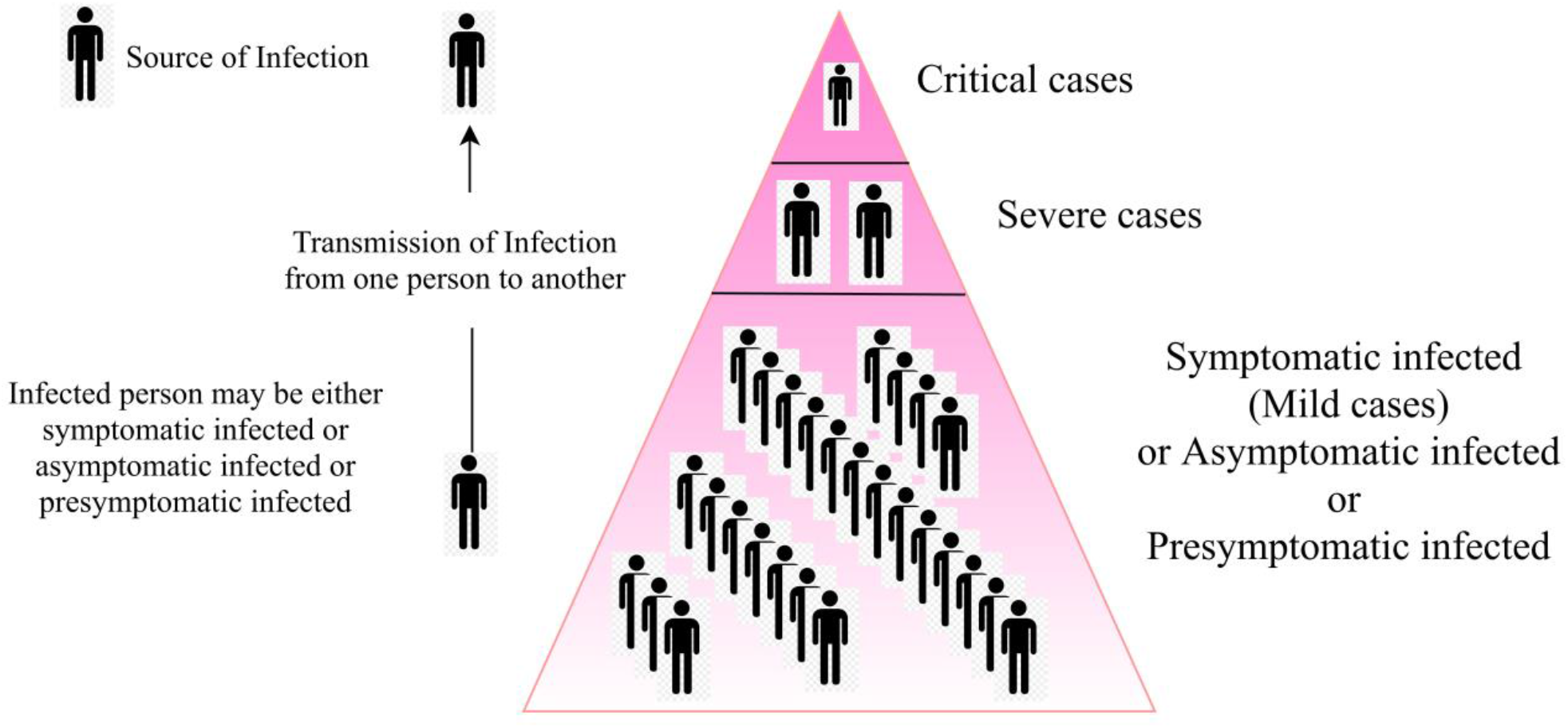
Hierarchy of infection

The Directive issued by the public health department of Brihan Mumbai Municipal Corporation (BMC) [25] says, “All public and private hospitals are instructed that no asymptomatic COVID-19 positive patient shall be given admission to ensure prompt availability of beds for the needy and genuinely deserving symptomatic patients. All asymptomatic patients admitted to various public and private hospitals shall be urgently discharged.

COVID-19 Infectious disease is classified mainly as symptomatic infectious and asymptomatic or pre-symptomatic infectious. Fig.5 indicates the categorization and flow of infectious events and cases and symptoms of mild cases quarantine in the home and severe and critical cases put in the hospital for proper treatment.

**Figure 5:**
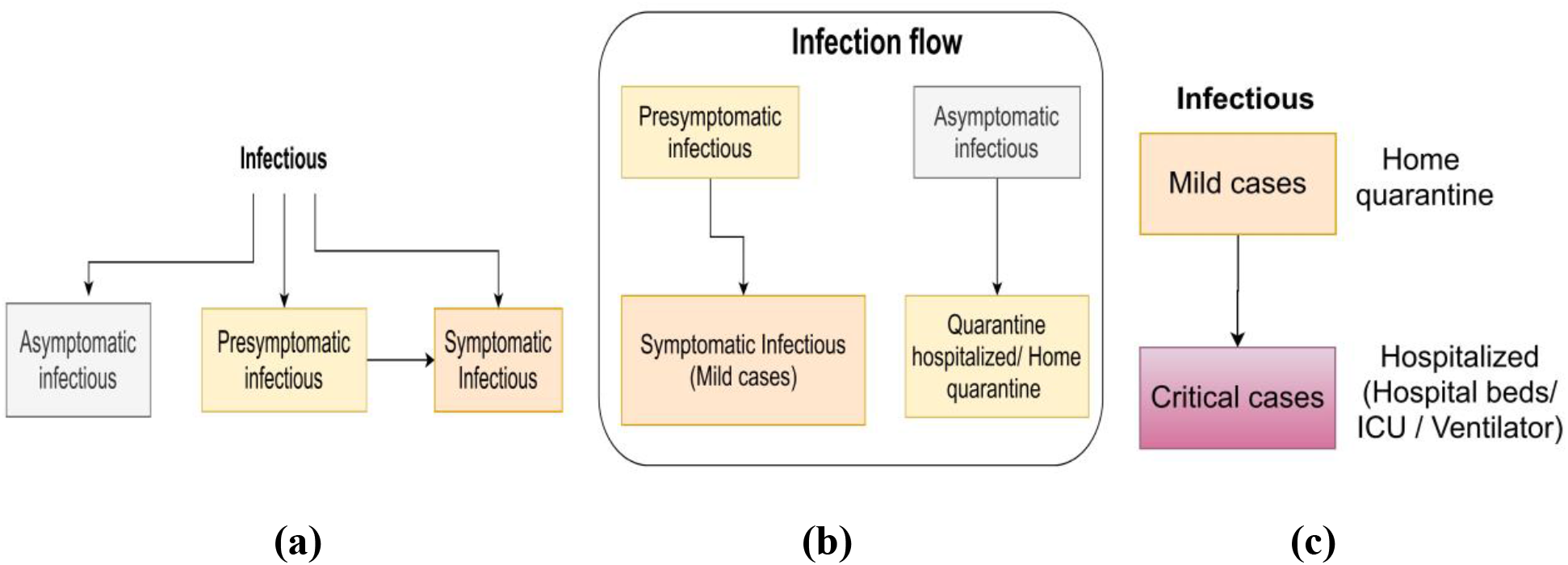
**(a)** Infectious cases categorization, **(b)** Infection flow, **(c)** Mild and severe infection flow

Fig.6 shows the flow of the proposed model. Initially, we consider all cases to be susceptible. The overall population is constant throughout the spread of infection. After exposure, some cases are found to be asymptomatic infectious with the help of contact tracing. Some cases are found to be presymptomatic infectious, in which the complete symptoms of the disease begin to appear within a few days. Some of the traced asymptomatic infectious conditions are severe, and then put into quarantine hospitalized after they have recovered. In the case of Symptomatic Infectious, most cases are mild, and some are severe and critical when taken to the hospital for treatment. After treatment, some cases will be recovered, and some will be dead due to infection.

**Figure 6:**
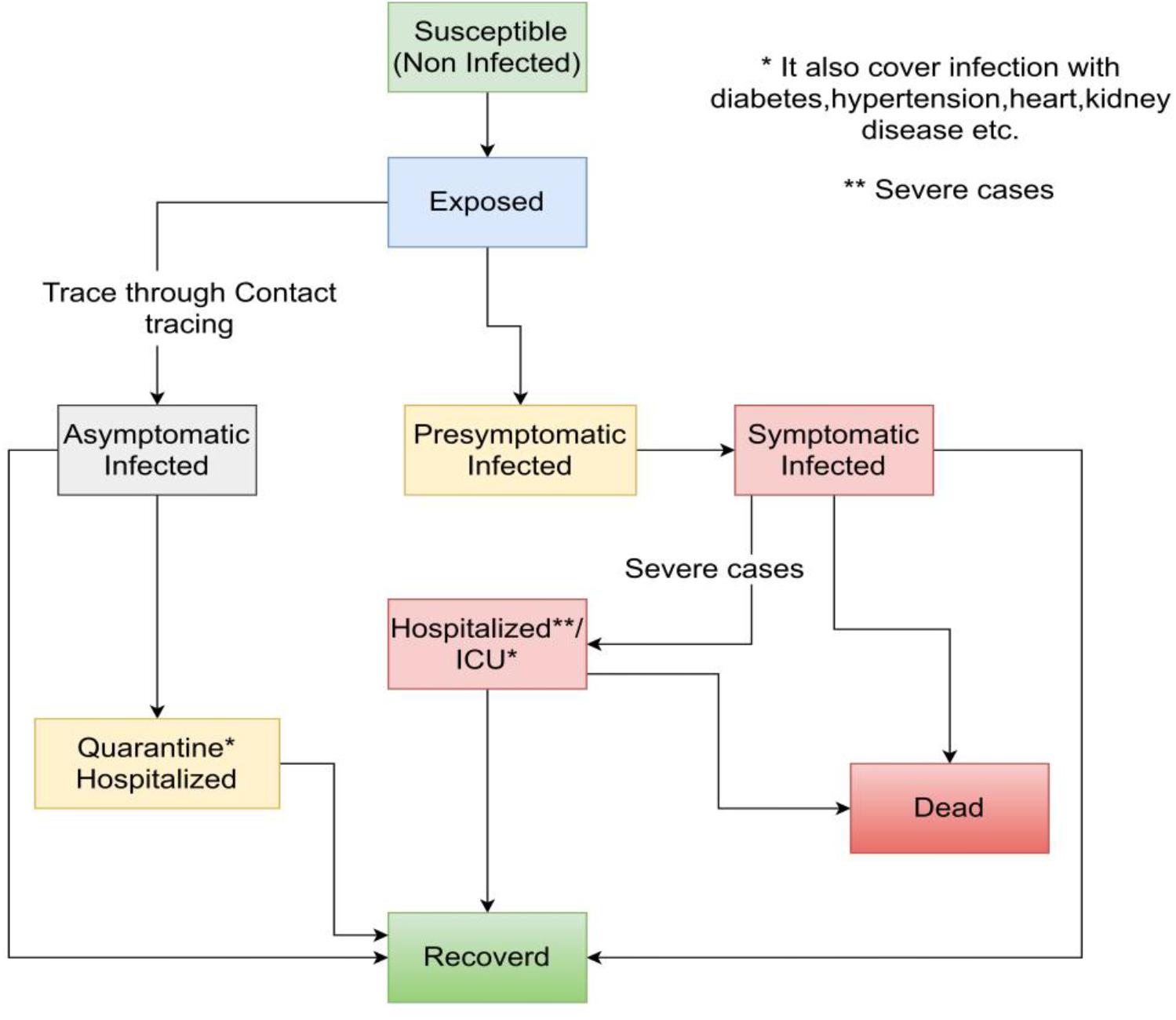
Flow of proposed model

Compartment models like SIR and SEIR are mostly used today for the mathematical modelling of infectious diseases. The basic compartmental SIR model has been proposed by Kermack-McKendrick [26] in 1927. Cooke and Driessche [27] studied the incubation cycle in the propagation of infectious diseases, developed the ‘Exposed, E’ compartment, and suggested an SEIR model of time delay. The basic compartment model SIR, the model SEIR, and the model SEIRD has been described by Singh A et al. (2020) [6].

In this work, we use a compartmental differential equation model for the propagation of COVID-19 in Mumbai. The model tracks the dynamics of the nine sub-population compartments, which are susceptible (S(t)), exposed (E(t)), asymptomatic infectious (A(t)), presymptomatic infectious (P(t)), quarantine hospitalized (Q(t)), symptomatic infectious (I(t)), hospitalized/ICU (H(t)), recovered (R(t)), and dead (D(t)).

The model simulations will be carried out with the following assumptions:

a. We are doing an age-wise study of spread scenarios.
b. The susceptible and infected individuals are homogeneous in the population.
c. No vaccine has been used with this model to stop the spread of COVID-19.
d. The population is constant. In our model, no recruiting individuals are permitted.

### Susceptible compartment S(t)

This compartment’s population will remain constant because no recruiting individuals are allowed in our model. This compartment population will decrease after infection due to an interaction with an infected person or an asymptomatic person.

### Exposed compartment E(t)

**This compartment’s population** consists of infected individuals but cannot infect others, and the population decreases with rate δ to become presymptomatic infectious or asymptomatic infectious.

### Asymptomatic infectious compartment A(t)

This compartment population is considered infected, but asymptomatic individuals do not develop common symptoms of COVID-19. Asymptomatic individuals are essential to model because they have the ability to spread the virus without knowing it, and with the help of tracing, we find these individuals.

### Presymptomatic infectious compartment P(t)

The population of this compartment is considered as infected, who have not developed symptoms yet, but who later develop symptoms.

### Symptomatic infectious compartment I(t)

The population of this compartment is considered as infected. Symptomatic infected individuals exhibit a common symptom of COVID-19. When symptomatic infected individual conditions are critical, they are admitted to the hospital for treatment. Some cases have been recovered, and some cases have been confirmed to be dead.

### Quarantine hospitalized compartment Q(t)

Older people. Asymptomatic patients who do not have a chronic disease do not need to be hospitalized and may opt for quarantine hospitals if hospitalization is necessary.

### Recovered compartment R(t)

Recovered cases are referred to as individuals who have gained immunity from COVID-19 disease. We have assumed that the cases which have been recovered will be immunized.

### Dead compartment D(t)

All infected individuals who died of COVID-19 disease come under this compartment.

We are consider that *N* = *S* + *E* + *A* + P + *I* + Q + H + R + *D* is constant, where *N* is the size of the population modeled. Fig. 7 shows the proposed model.

**Figure 7:**
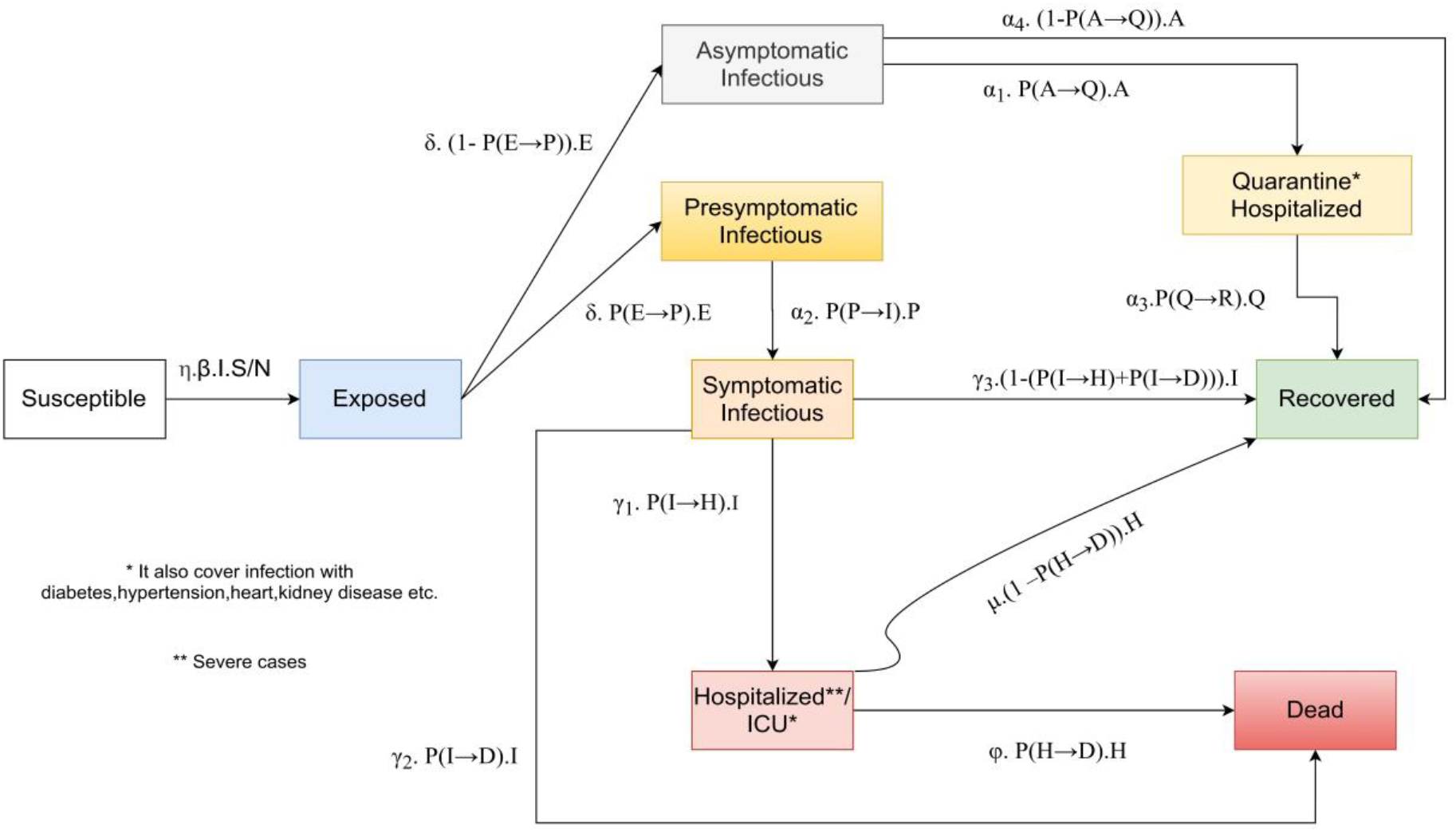
Proposed model

#### Differential Equations for proposed model

The ordinary differential equations of the proposed model are as follows:

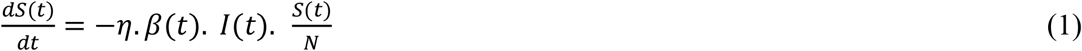

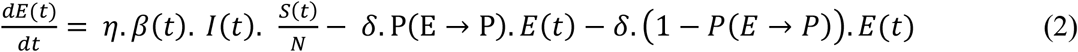

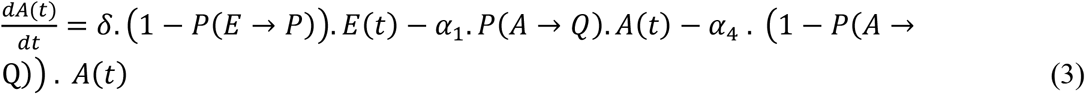

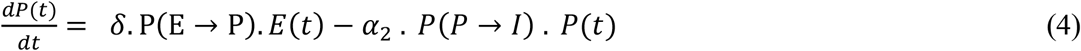

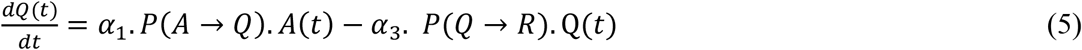

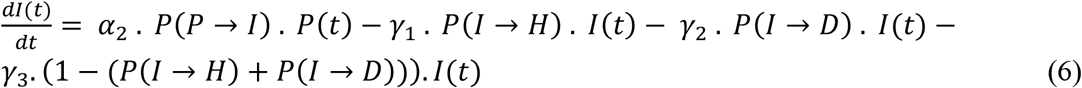

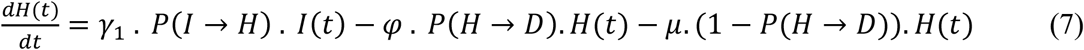

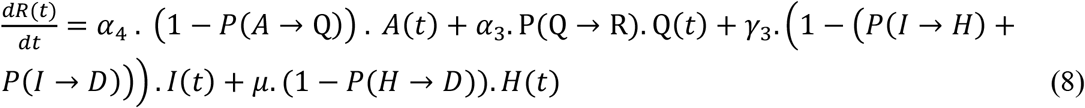

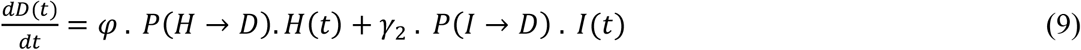

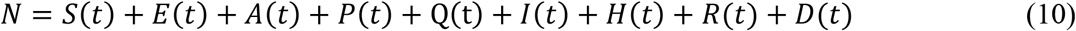

All parameters with description:

- *η* – Social distancing factor
- *β* – Rate of transmission
- δ – The rate of infection transmission from exposed to Presymptomatic infectious or Asymptomatic infectious
- α_1_ –Median time from Severe Asymptomatic Infectious to admission in Quarantine Hospital
- α_2_ – Median time from Presymptomatic to Symptomatic infectious
- α_3_ – Median time to recover from Quarantine Hospital
- α_4_ –Median recovery time of Asymptomatic infectious
- γ_1_ – Median time for developing pneumonia and other hospitalization symptoms
- γ_2_ – Median time from symptomatic infection (without hospitalization) to death
- γ_3_ – Median recovery time of Symptomatic infectious
- φ – Median time to stay at the hospital/ICU
- μ – The time of recovery for hospitalized people

There is no known exact value of the initial conditions (S(0), I(0), R(0)). We put very carefully the value of the initial conditions, because these ordinary differential equation systems are highly sensitive to initial parameters.

First, we fit the available clinical data, and then we estimate the model parameters values from it. We used the least square method and the Levenberg-Marquardt model to calculate the parameters R_0__start, R_0__end, k, x_0_, s, P(E→P), P(A→Q), P(Q→R), P(P→I), P(I→H), P(H→D) and P(I→D) for the proposed model. To estimate the final value of any parameter, we first set the range of the parameter means the maximum value and the minimum value. Then we have to set the initial value of the parameter to fit the model and get the final fitted value. We do not need to fit some parameter, for example, η, δ, α_1_, α_2_, α_3_, α_4_, γ_1_, γ_2_, γ_3_, φ, μ etc. need not to fit. We have to get these parameter values from the review of research papers and reports of trusted organizations. Tab. 2 and Tab. 3 shows the numerical values of the model parameters for selected countries.

**Table 2:** Value of parameters from authentic sources

| Parameters | Description | Value |
| --- | --- | --- |
| $\eta$ | Stringency index | 1 |
| $\delta$ | Exposed to Presymptomatic / Asymptomatic infectious | 1/3.5 |
| $\alpha_1$ | Asymptomatic to Quarantine hospitalized | 1/2.0 |
| $\alpha_2$ | Presymptomatic to Symptomatic infectious | 1/2.0 |
| $\alpha_3$ | Quarantine hospitalized to Recovered | 1/10.0 |
| $\alpha_4$ | Asymptomatic infectious to Recovered | 1/2.0 |
| $\gamma_1$ | Symptomatic infectious to Hospitalized | 1/3.7 |
| $\gamma_2$ | Symptomatic infectious to Dead | 1/6.4 |
| $\gamma_3$ | Symptomatic infectious to Recovered | 1/10.0 |
| $\varphi$ | Hospitalized to Dead | 1/2.4 |
| $\mu$ | Hospitalized to Recovered | 1/10.0 |

**Table 3:** Parameters estimation with fitted value

| Parameters | Initial Value | Minimum Value | Maximum value | Fitted Value |
| --- | --- | --- | --- | --- |
| P(E→P) | 0.3 | 0.2 | 0.5 | 0.2 |
| P(A→Q) | 0.9 | 0.85 | 1.0 | 0.900438516 |
| P(Q→R) | 0.9 | 0.8 | 1.0 | 0.999642426 |
| P(P→I) | 0.9 | 0.85 | 1.0 | 0.874977863 |
| P(I→H) | 0.1 | 0.05 | 0.25 | 0.239551274 |
| P(H→D) | 0.25 | 0.20 | 0.70 | 0.451287751 |
| P(I→D) | 0.025 | 0.01 | 0.10 | 0.067009916 |
| s | 0.003 | 0.001 | 0.01 | 0.003141659 |
| R_0_start | 1.5 | 1 | 2.2 | 1.51633527 |
| k | 0.2 | 0.1 | 0.8 | 0.2 |
| x0 | 180 | 0 | 380 | 180 |
| R_0_end | 0.4 | 0.3 | 1.0 | 0.4 |

The proposed method mainly focused on death cases because death cases hardly go undetected. The proposed methods result shows the predicted outcome is very closed to the real scenario.

## 3. Data Analysis and parameter estimation

Mathematical modeling of infectious diseases is simplified by compartmental models. Compartmental models are based on ordinary differential equations. To solve these ordinary differential equations, some initial conditions and characteristic parameters are needed. It is very sensitive to these initial parameters. A minor change in them can make a big difference in results, so in the equations they need to be carefully implemented. Preliminary parameter estimation helps estimate the case fatality rate and basic reproduction number. The social distance parameter suggests that social contacts and physical contact would be avoided to control the transmission of infectious diseases. The meaning of the social distancing factor lies between zero and one. When a lock is applied, and all individuals are placed in quarantine, its value is zero, and if they obey unregulated and routine, its value is one.

### The basic reproduction number R_0_ and case fatality rate over time

The ratio between the transmission rate and the recovery rate is the basic reproduction number R_0_. The basic reproduction number is the average number of people infected by any an infected person, if no intervention measures are in place. The basic reproduction number is important for disease intervention. The value of the basic reproduction number R_0_ varies over time; the basic reproduction number value decreases if any country embraces lock-downs, and its value starts to increase again when that country eliminates the lock-down.

R_0_ is the ratio of β and γ in the compartmental models (R_0_=β/γ). Hence, we can say that *β* = *R*_0._ *γ*. In this manuscript, before the lock-down, the value of R_0_ is R_0_ start, and if the lock-down is imposed on day L, the value of R_0_ is reduced to R_0_ end, and the value of β will be changed accordingly. In real life, the basic reproduction number R_0_ is not constant and changes over time. When social distancing is imposed or removed, its value will change accordingly. The model illustrates the initial impact of social distancing on the basic reproduction number. The time-dependent variation in R_0_ can be given as

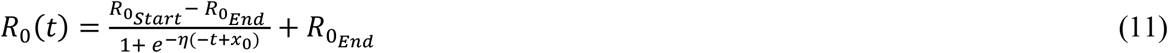

The description of the parameters is given below:

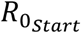 – The value of the R_0_ for the first day

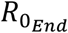 - The value of R_0_ for the last day x_0_ is the day when R0 drastically decreases or inflection point

η is the parameter of social distance.

The value of the x_0_ should be given very carefully because it makes a big difference to the result and the value of the 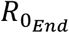 must be less than one.

### Age-dependent fatality rate

The infection mortality rate is the ratio between the number of casualties and the number of cases. The rate of death is not constant and depends on several variables such as age, fatal diseases, etc. If the majority of people are infected, then the fatality rate is high, and the fatality rate is low when fewer people are infected. If infected people are very high, we need more health care. Often, not everyone is treated because of the shortage of medical services. We need, therefore to know what proportions of individuals are currently infected. Therefore we describe the fatality rate φ as

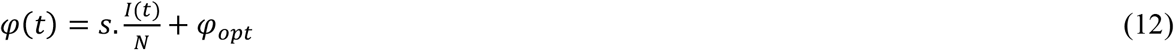

The description of the parameters is given below:

*s* - It is arbitrary, but the fixed value that controls infection

φ _opt_ - optimal fatality rate

As it governs the above equation, the value of parameter s is significant; very small changes in s can make a huge difference in performance. The value of this parameter should therefore be carefully chosen. At time t, I(t) is the sick, and N is the total population.

Depending on the age group, case fatality analysis is complex. Therefore, to help assess the case fatality, we classify age into various age classes (e.g. people aged 0 to 9, aged 10-19,….., aged 90-100). The fatality rate by age group and the proportion of the total population in that age group are two things that are needed for the age group study. If the elderly population is higher, the death rate will rise, and if the proportion of young people is higher, the death rate will be lower.

Therefore, the Death (fatality) rate is calculated as follows:

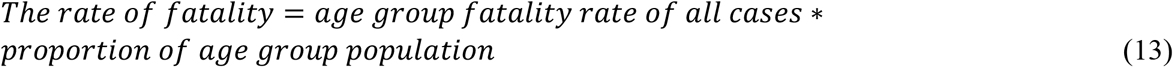

The ratio between the number of deaths and the number of cases is the death rate. The risk of dying can be described as being infected by a virus (in percentage) and depends on different age groups. The following Table 4 presents the percentage of deaths in a given age group if a person is infected with COVID-19 [32].

**Table 4:** Age-wise death rate in all cases

| Age Group | Death Rate |
| --- | --- |
| 90+ years old | 8.60% |
| 80-89 year old | 20.89% |
| 70-79 years old | 20.40% |
| 60-69 years old | 29.28% |
| 50-59 years old | 23.75% |
| 40-49 years old | 11.37% |
| 30-39 years old | 4.59% |
| 20-29 years old | 1.56% |
| 10-19 years old | 0.32% |
| 0-9 years old | 0.24% |

### Hospitalized cases analysis

We know every country has a limited number of hospital beds. In order to raise the number of hospital beds when the disease spreads, all countries are starting new hospitals, opening rooms, etc. The number of hospital beds is therefore, increasing over time. We can model the number of beds as follows:

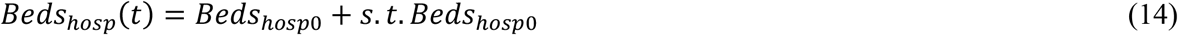

Description of the parameters is given below:

Beds_hosp0_ - Total number of Hospital beds available

*s* - some scaling factor,

Since the number of beds displayed in the formula varies every day to decide how many beds has been served every day in hospitals for patients, this is a significant factor.

### Environment setting

In this study, Anaconda development platform are considered for running the proposed model. The PYTHON platform used some important libraries such as pandas, NumPy, LMfit, ode, etc. for all the experimental studies. The hardware configuration of the system are reported in Table 5.

**Table 5:** Specification of System

| Name | Parameter |
| --- | --- |
| System RAM | 12 GB |
| The processor | Intel(R) Core (TM) i7-5500U CPU |
| The processor Speed | 2.40 GHz |
| Cores | 2 Core(s) |
| Logical Processor | 4 |
| GPU RAM | 4 GB |
| Graphics processor | NVIDIA GeForce GTX 950M |
| Cuda cores | 640 |
| Memory interface 256-bit | 128-bit |
| Memory type | DDR3 |
| Bandwidth 288.5 GB/s | 28.80 GB/s |
| Language | Python |

### Fitting the model to find some important parameters value

In this manuscript, we concentrate on fitting a model with the basic reproduction number and age group fatality, hospital beds with actual COVID-19 data, which are close to the real data and find parameters that can be useful in future discussions for our model. The initial assumptions of the parameters are extremely important. First we need to understand what parameters are understood and what we need to get out of them. Many important parameters have been used in this proposed model. Some of the parameters are calculated and considered by the current study and data. It’s not necessary to fit N, put the total population count in place of N. According to the current data and analysis, we value parameters, i.e. social distance parameter β, in the range 0-1 where 0 shows everyone is in lock down and quarantined, while one is for a normal way of life.

First from reliable sources, we collected all the related COVID-19 data and parameters available to us and then defined initial guesses and limits within certain limits, depending on the situation, for the remaining parameters of this model. To extract different parameters from the data fitting, we have used the Levenberg-Marquard algorithm, as this is the most significant and effective data fitting algorithm reported[33, 34]. For Python, the LMfit library provides a high quality interface for non-linear optimization and curve-fitting issues[35]. The proximity of the data to the fitted line is measured by the R-squared value. The R-square value is between zero and one at all times. Usually, the higher the R-squared value, the better the model that your data is given. Table 6 shows the R-squared value of Mumbai for death cases. The curve of Mumbai death cases is shown in Fig. 8.

**Table 6:** R-squared value of curve fitting for Mumbai

| City | R-squared value (Death cases) |
| --- | --- |
| Mumbai | .972 |

**Figure 8:**
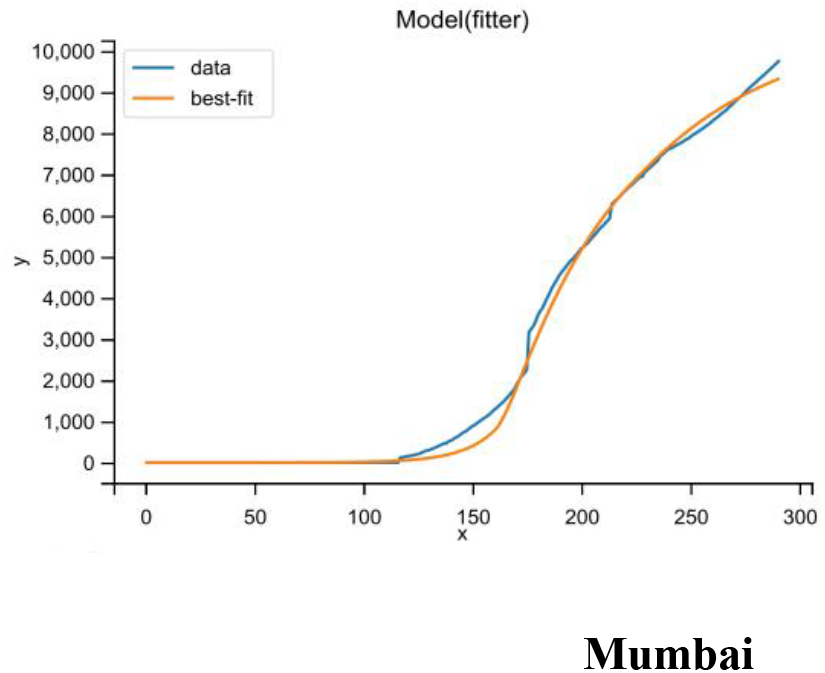
Curve fitting of death cases in Mumbai

The main source of data is www.covidindia.org [29]. For age groups, probability, and hospital beds, we have collected and data from the Indian government data center [28]. From January 30, 2020 onwards, time series data are available. These data are collected through public health authorities with ethical permission.

#### Distribution of Total cases worldwide

According to WHO, 216 countries are affected by COVID-19. Fig. 9 (a) shows that 15% of cases come from India. In comparison with the number of global total cases, Mumbai shares one percent.

**Figure 9:**
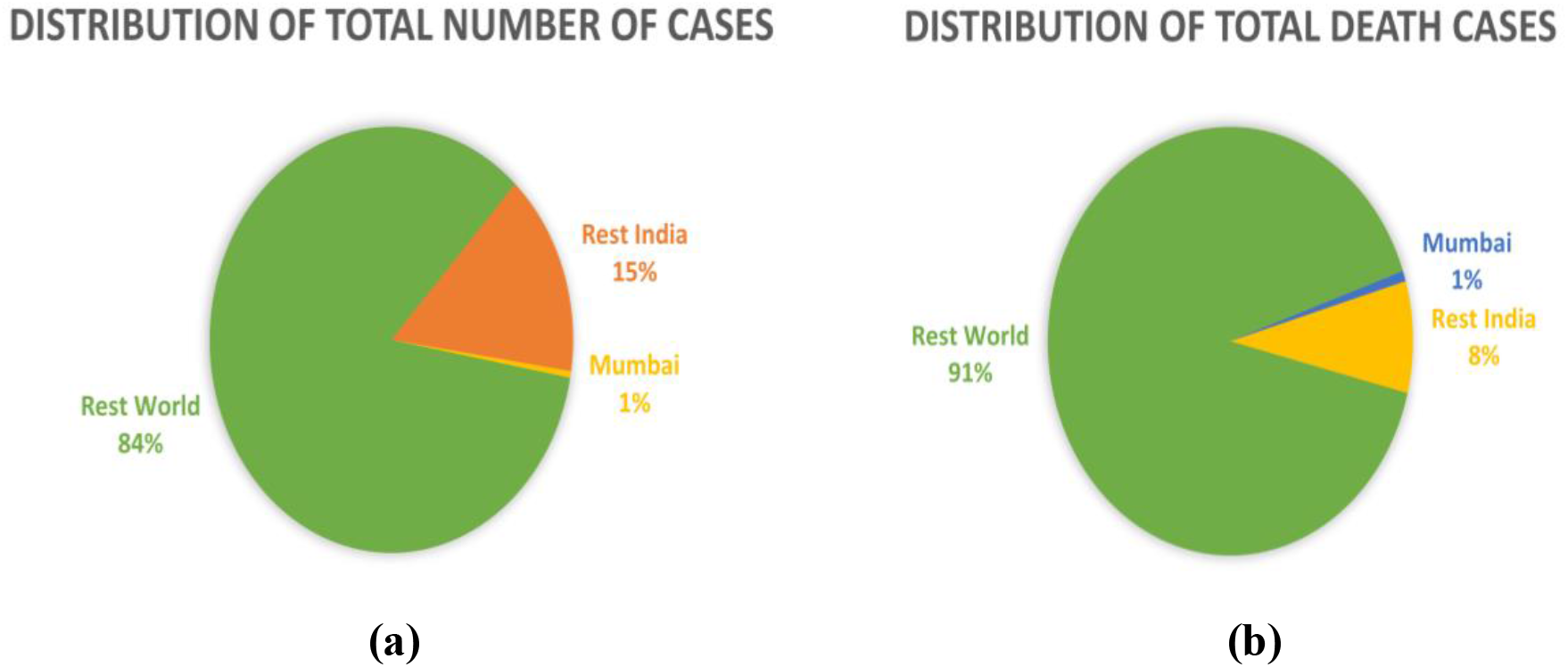
**(a)** Distribution of total cases worldwide, **(b)** Distribution of Death cases worldwide

#### Distribution of Death cases worldwide

Death cases are more accurate because the associated authorities rarely miss report of death due to COVID-19; hence, death cases are more accurate. Fig. 9 (b) shows that 8% of death cases come from India. In comparison with the number of global total death cases, Mumbai shares one percent.

#### Day wise total cases and total death cases

The Fig. 10(a) shows the total cases and Fig. 10(b) shows the death cases of Mumbai from January 30, 2020 to September 30, 2020 of COVID-19.

**Figure 10:**
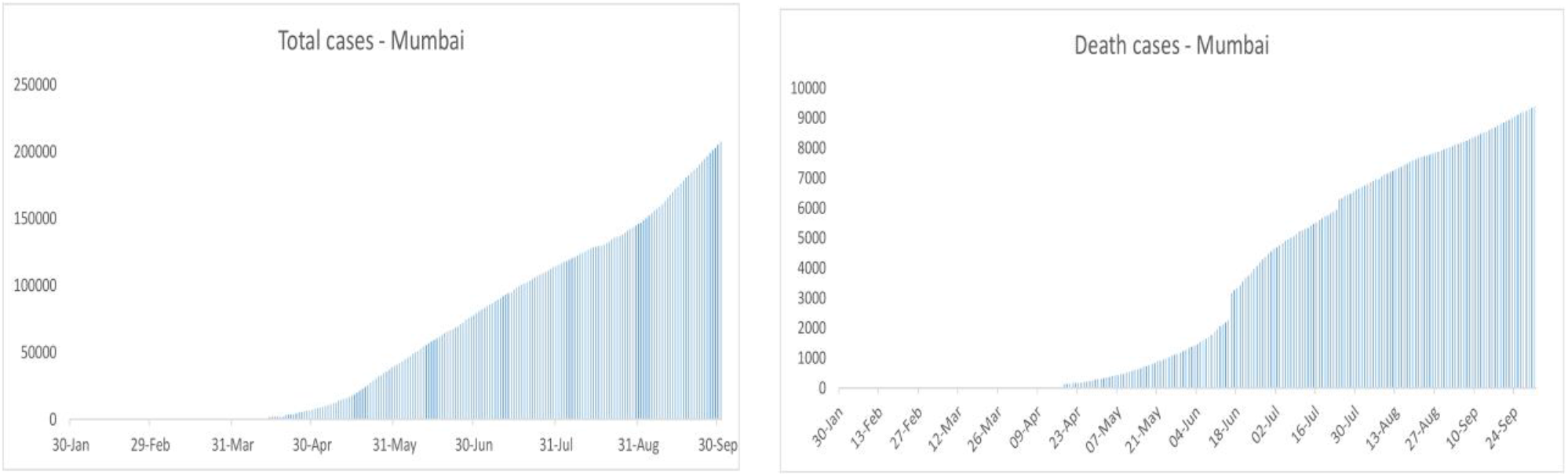
**(a)** Day-wise total cases of Mumbai, **(b)** Day-wise total death of Mumbai

#### Daily new cases and daily death cases

Fig. 11 (a) shows the daily new cases and Fig. 11(b) shows daily new death of Mumbai from January 30, 2020 to September 30, 2020 of COVID-19.

**Figure 11:**
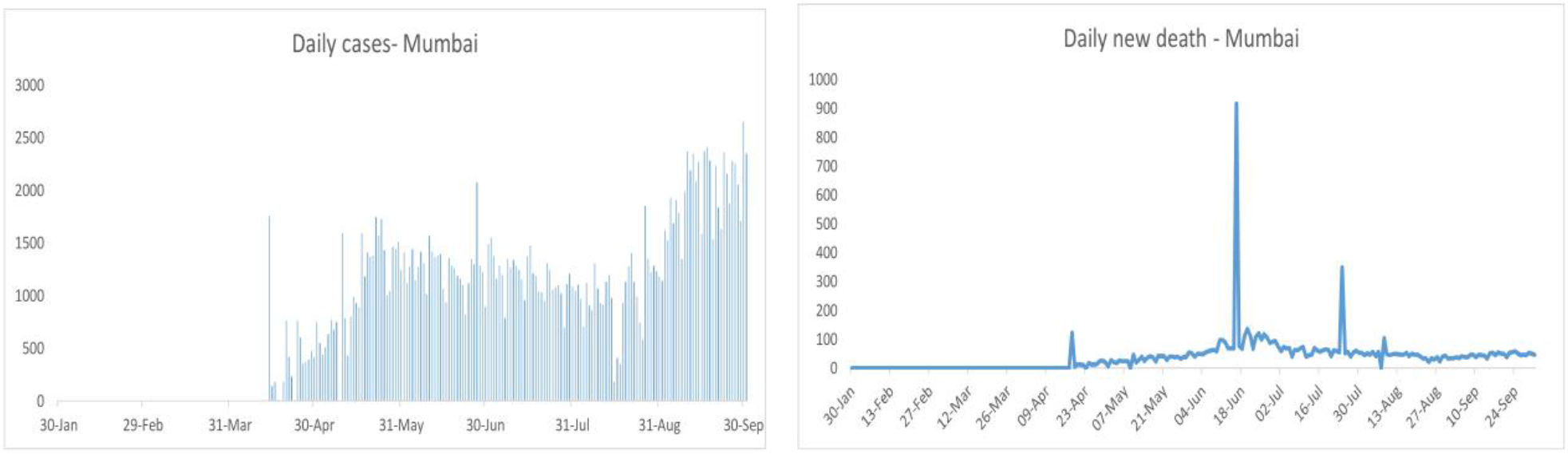
**(a)** Daily new cases of Mumbai, **(b)** Daily new death cases of Mumbai

### Age-wise population

In terms of age, the majority of the population in India is young. The population of old age on this continent is much smaller. The population distribution by age group is represented in Fig. 12 [31].

**Figure 12:**
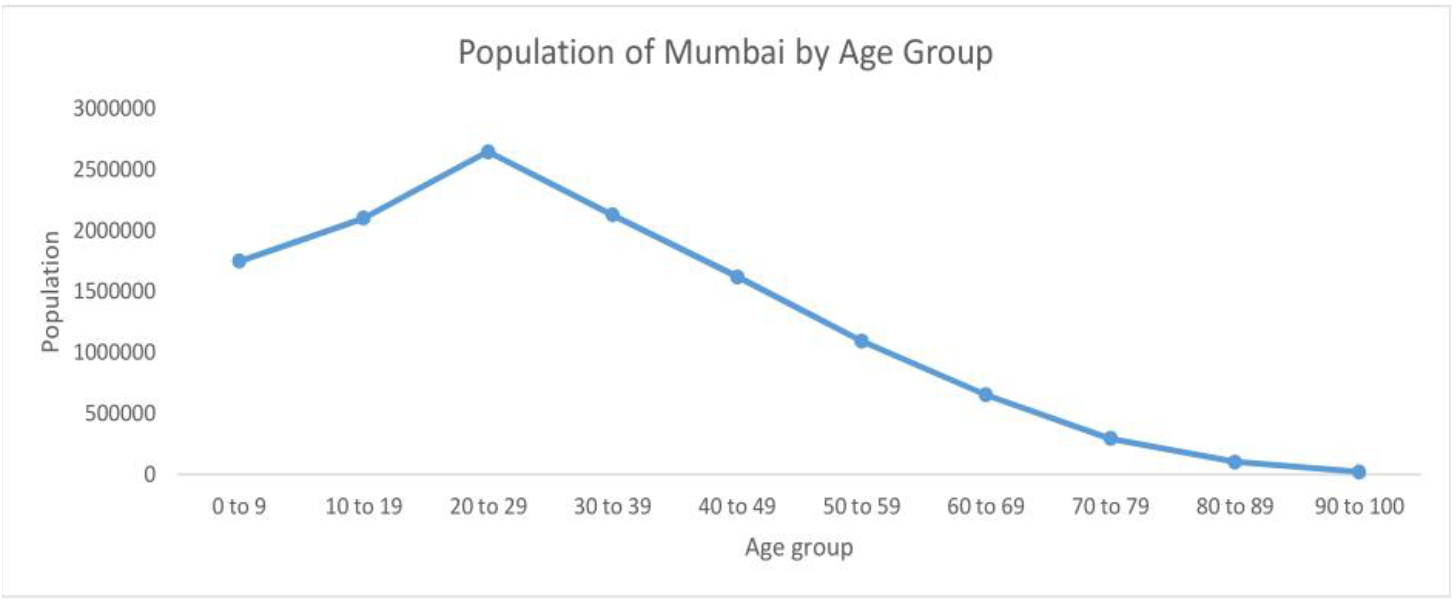
The distribution of the population according to the age category of Mumbai

#### The number of hospital beds per 100k

Unit beds per 100,000 population or a 100k population, according to WHO and the OECD [30]. Basic measures focus on all hospital beds that are occupied or empty. Mumbai city number of hospital beds is shown in below Table 7:

**Table 7:** As of 13 October condition of hospital beds and quarantine beds

| Types of Bed | Bed Capacity | Occupied | Available |
| --- | --- | --- | --- |
| DCH & DCHC Bed | 15057 | 9827 | 5230 |
| ICU Bed | 1998 | 1725 | 273 |
| O<sub>2</sub> Bed | 9145 | 5869 | 3256 |
| Ventilator Bed | 1154 | 1032 | 122 |
| CCC1 Facilities* | 46677 | 1047 | 45630 |
| CCC2 Facilities* | 23806 | 1150 | 22656 |

\* Quarantine Hospitalized

## 4. Results and Discussion

We analyze and forecast in this section, using our proposed model, the pattern of COVID-19 of Mumbai. The result of our proposed method shows that Mumbai have experienced their worst time. In Mumbai, the distribution of disease is very random, e.g., the number of cases in Wards RC, RS,KW,PN,KE, KE, GN, S, and Ward N is very high as compared to other Wards. If we compare the results of our proposed method with real data, it is approximately close. In this section, we calculate for Mumbai the basic reproduction number, the case fatality rate, and the number of hospital beds required at peak time.

### 4.1 Proposed model for Mumbai

Mumbai is the most affected country by the COVID-19 disease. According to the proposed method, cases in Mumbai will start to decline continuously after the last week of September, and the cases will end approximately in the last week of March. The cases reach on the peak in Mumbai, according to our proposed method, in September and October. The results of our proposed method show that there will no need of Hospital beds in near future. The result shows the basic reproduction rate is 1.8 in starting, and after some time, it starts decreasing visually and goes up to one and even lower. Proposed model for the Mumbai shows that the case fatality rate of COVID-19 in Mumbai is moderate. There are many up2s and downs in the daily case fatality rate, but it goes higher and higher up to 5.5 in our model and then slows down, and total CFR is nearly 4.6. During peak time, the number of death cases is 65 per day, and daily-hospitalized cases are nearly 350. Proposed model results for the Mumbai has shown in Fig. 13.

**Figure 13:**
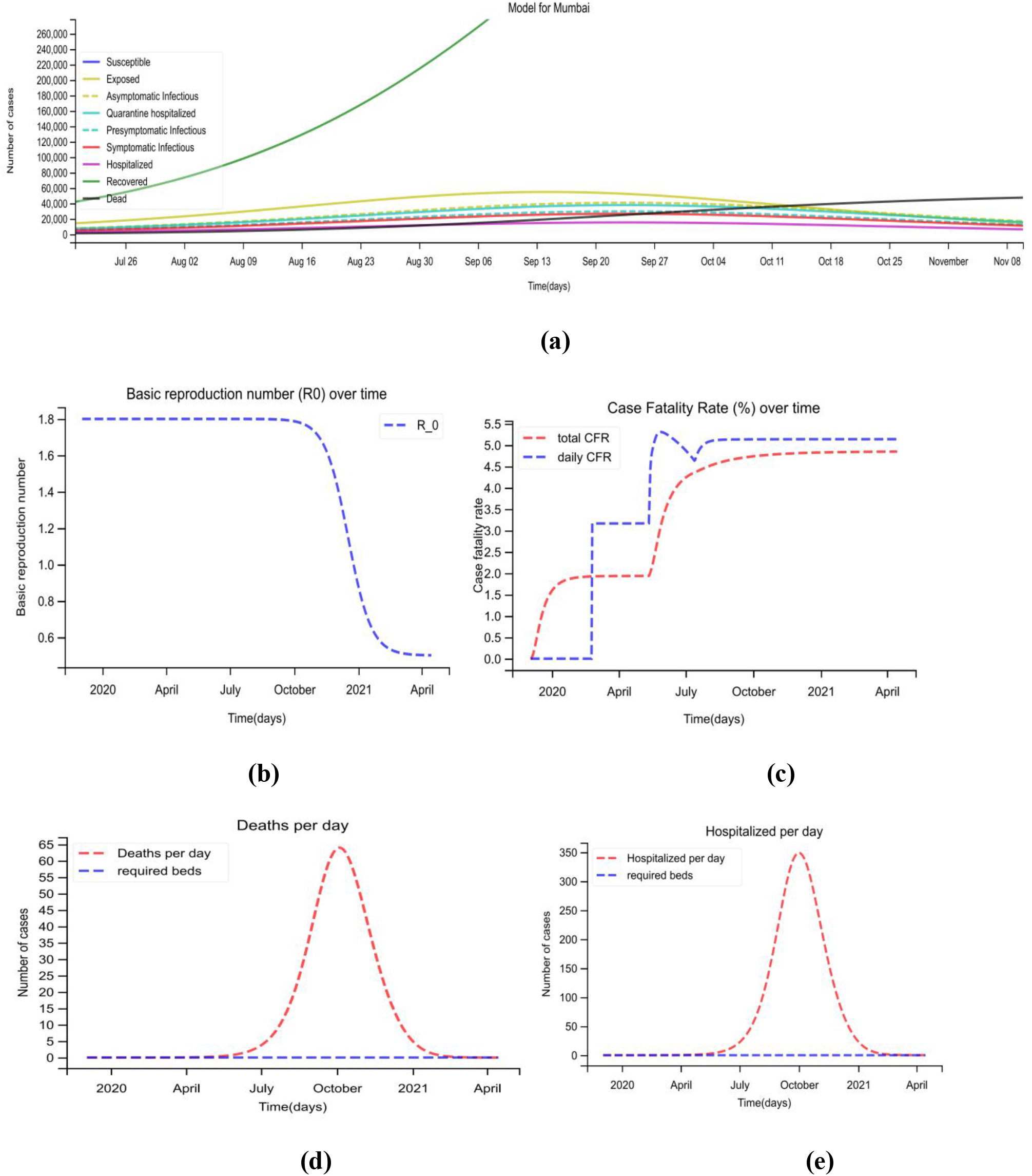
Proposed model for Mumbai. **(a)** A 500-day analysis has been done by the proposed model, which starts from January 30. **(b)** The basic reproduction number of Mumbai over time **(c)** the case fatality rate of Mumbai over time. **(d)** Daily death cases in the Mumbai, where the red line shows the number of deaths per day and the blue line shows how many hospital beds are required during peak days. **(e)** Mumbai Daily Hospitalized cases, where the red line indicates the number of hospitalized cases per day, and the blue line shows how many hospital beds are required during peak days.

Fig. 14 shows the comparison between actual and forecast data for active cases with 95% interval. Rarely some of the forecast data go beyond the interval decided. For validation purpose, we predict the data for 25 days. Fig. 15 shows the comparison between actual and forecast data for daily new cases with an interval of 95%, and Fig. 16 shows the comparison between actual and forecast data for daily death cases with an interval of 95%. Confidence interval is basically the sensitivity analysis to determine the robustness of model predictions to parameter values (Reproduction number) and the sensitive parameters are estimated from the real data on the COVID-19 pandemic in Mumbai.

**Figure 14:**
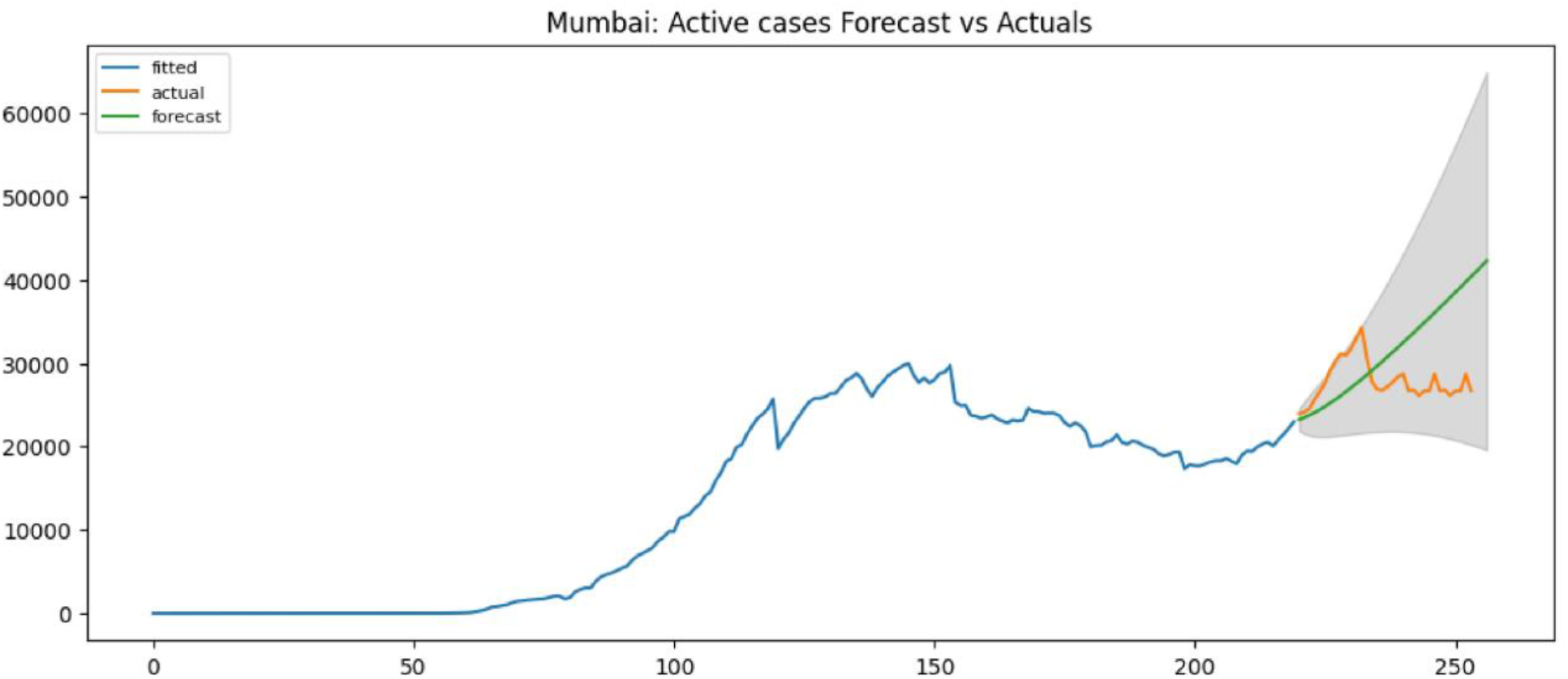
Compare the actual and forecast active case data with 95% interval

**Figure 15:**
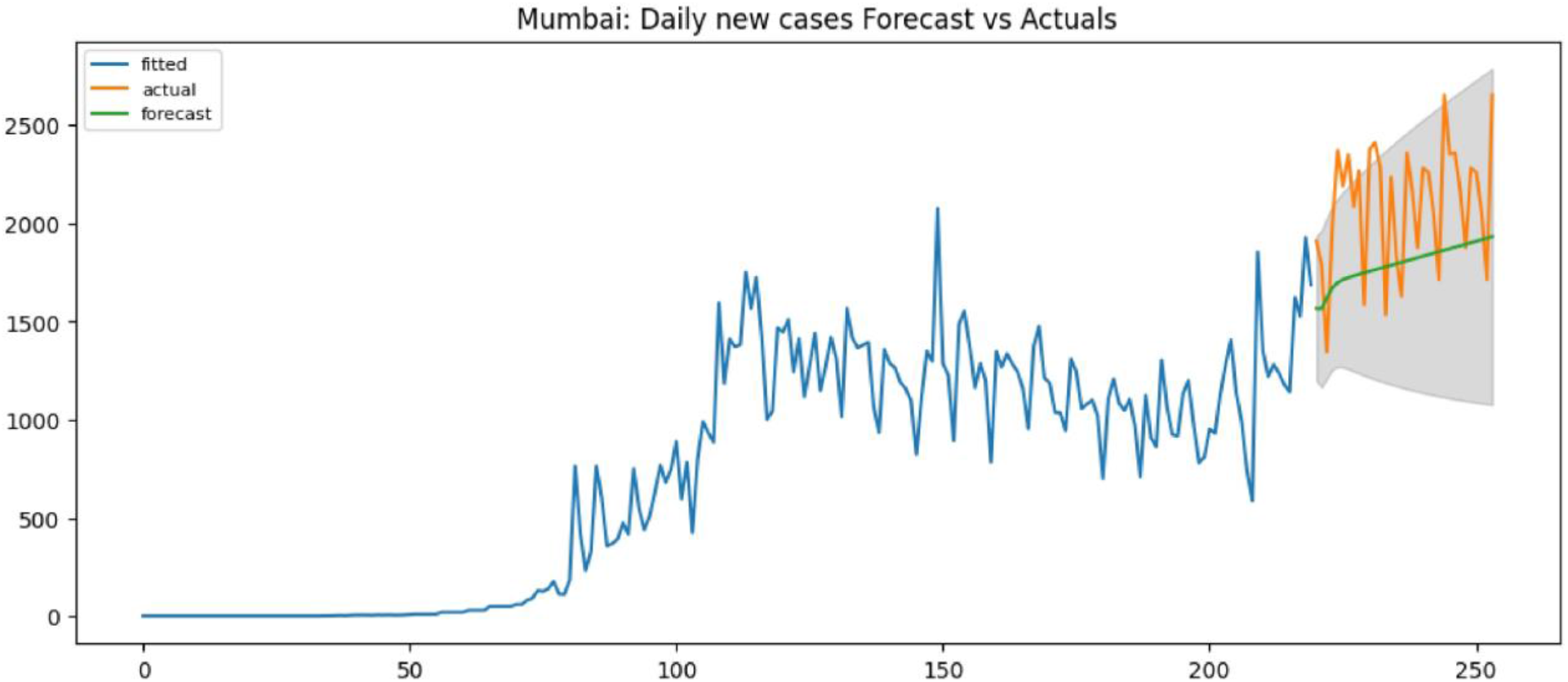
Compare actual and forecast data for daily new cases with an interval of 95%

**Figure 16:**
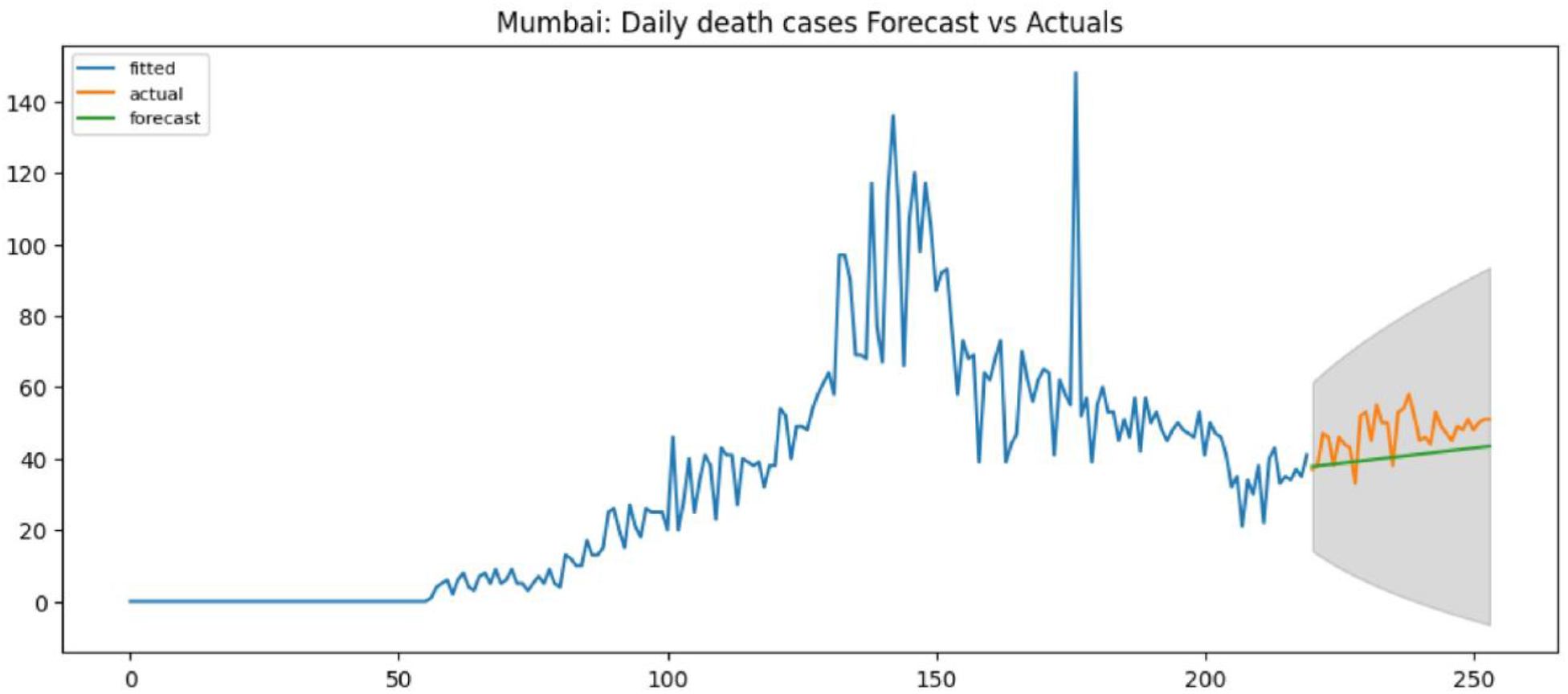
Compare actual and forecast data for daily death cases with an interval of 95%

The basic reproduction number for COVID-19 initially 1.8, but later it goes up to 0.5. After October 2020, the spread of disease decreasing. The summary of the proposed model results for the selected countries is shown in Tab. 8.

**Table 8:** Summary of Infection Propagation for Mumbai

| Description | Mumbai |
| --- | --- |
| Approximate time when the disease is on peak | 180-250 |
| The approximate time when cases of death rarely occur | 320 |
| Basic reproduction number (highest) | 1.8 |
| Case fatality rate (highest) | 5.1 |
| Overall case fatality rate | 4.6 |
| Approximately needed Hospital beds while the disease is at a peak | No |
| Approximately the needed ICU beds when the disease is at a peak | No |
| Highest number of people dead in a single day | 65 |
| Highest number of people hospitalized in a single day | 350 |

### 4.2 Performance Evaluation criteria

The prediction performance of the proposed system is evaluated using the following metrics:

#### 4.2.1 Mean absolute percentage error (MAPE)

Analyze the efficiency of the forecasting model of our method, we use the mean absolute percentage error (MAPE) [36] or Mean Absolute Percentage Deviation (MADE) as the criteria standard. Its formula is express as the following equation

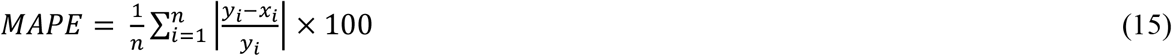

Where *y*_*i*_ denotes the *i*^*th*^ actual value, and *x*_*i*_ represents the *i*^*th*^ predicted value. If the value of MAPE is low, the accuracy of the method is high.

#### 4.2.2 Root Mean Square Error (RMSE)

Root Mean Square Error (RMSE) [37] or Root-Mean-Square Deviation (RMSD) is a measure of the average magnitude of the errors. Specifically, it is the square root of the average squared differences between the prediction and actual observations. Therefore, the RMSE will be more useful when large errors are particularly undesirable. If the value of RMSE is low, the accuracy of the method is high. RMSE formula is express as the following equation

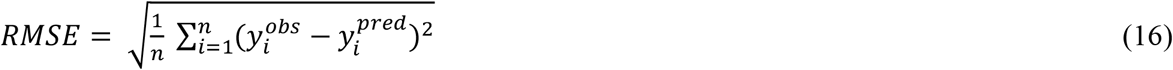

Where 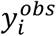 and 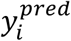 are the actual and predicted observations, respectively.

Our basic approach is to predict COVID**-**19 disease and analyze the spread scenario of COVID**-** 19 disease in India. Data used in the analysis are day wise sequential data. Hence if we need to compare our proposed work with deep learning models. LSTM (Long short**-**term memory) and Sequence**-**to**-**Sequence deep learning models are well suited for making predictions based on time series data.

The overall performance of deep learning methods is not better because there is less set of training data. If the training data set increases, the performance of the deep learning method will improve. For model validation, we have used precision measures, MAPE and RMSE. The proposed model outperforms the LSTM model and the Seq2Seq model. The result shows that the RMSE and MAPE accuracy of the proposed model better as compared to the LSTM and Seq2Seq models.

The performance evaluation compared with the following state of the art methods:

1. SIR (Susceptible Infectious Recovered Model)
2. ARIMA (Autoregressive integrated moving average Model)
3. SARIMAX (Seasonal ARIMAX Model)

The proposed model outperforms the state-of-the-art methods such as the SIR model, the ARIMA model and the SARIMAX model. The proposed model outperforms the LSTM model and the Seq2Seq model. The result shows that the RMSE and MAPE accuracy of the proposed model better as compared to the SIR, ARIMA and the SARIMAX models.

The results are evaluated using a variety of metrics (RMSE, MAPE) as well as graphically illustrated using Taylor diagrams [42–44]. Taylor diagram (Fig. 17) is used to perform the comparative assessment of several different models and to quantify the degree of correspondence between the modelled and observed behavior in terms of three statistics: Pearson correlation, RMSD (Root Mean Square Deviation), and the standard deviation. The model that is located lower on the diagram is considered to represent reality better.

**Figure 17:**
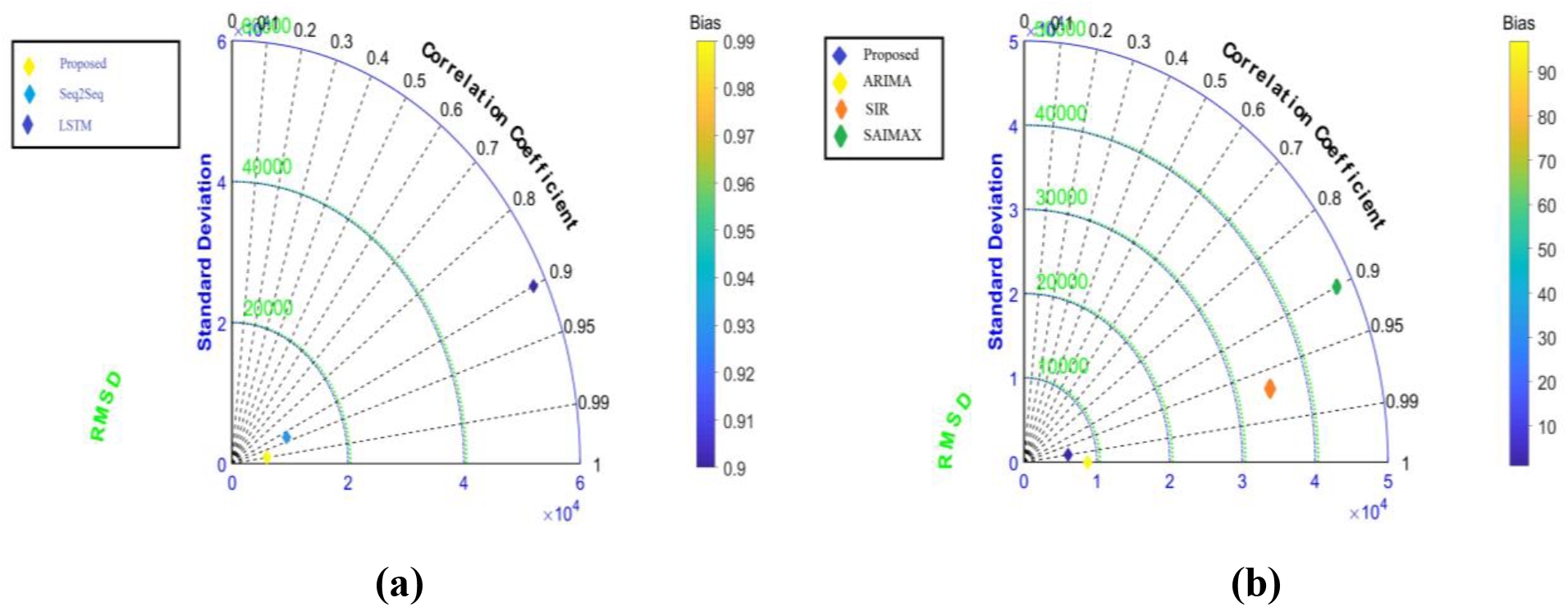
**(a)** Taylor diagram and proposed method compare with the deep learning prediction models, (**b)** Taylor diagram of proposed method compare with the state-of-the-art methods

Note that while the proposed model performs better than the ARX model and ARIMA model in terms of RMSD and total RSMD, it has a more considerable bias than the ARX model and ARIMA model. The results are summarized in Table 10. Table 9 summarized the LSTM, Seq2Seq model performances.

**Table 9:** Proposed method compare with the deep learning prediction models

| Prediction models | Root Mean Square Error (RMSE) | Mean Absolute Percentage Error (MAPE) |
| --- | --- | --- |
| LSTM<sup>1</sup>[38] | 57752.75 | 0.9363 |
| Seq2Seq<sup>2</sup>[39] | 10115.92 | 0.2497 |
| Proposed<sup>3</sup> | **6072.86** | **0.1836** |

LSTM<sup>1</sup> - Long short-term memory Model
Seq2Seq<sup>2</sup> - Sequence-to-Sequence Model
Proposed<sup>3</sup> - Proposed Method

**Table 10:**
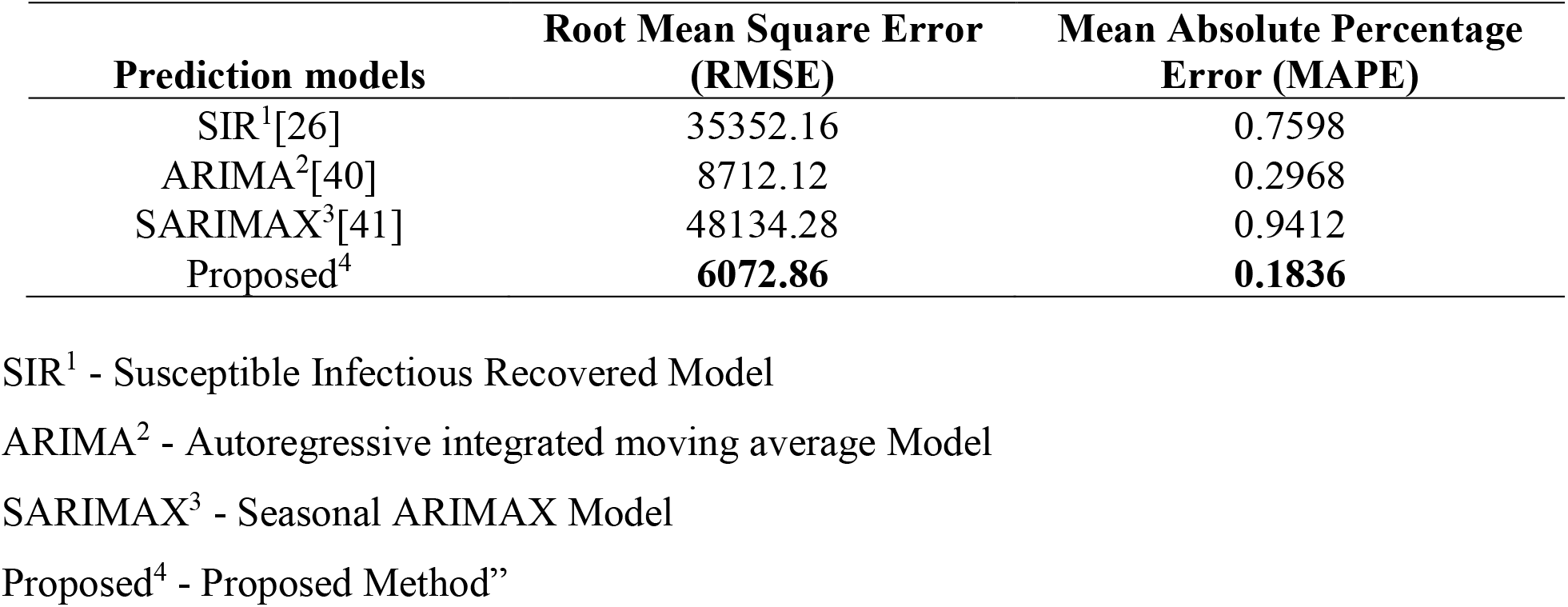
Proposed method compare with the state-of-the-art methods

## Discussion

Results from the proposed model suggest that this model has excellent potential for predicting the incidence of COVID-19 diseases in time series. The central aspect of our proposed model is the addition of hospitalized, quarantine hospitalized, presymptomatic and asymptomatic infectious compartment, which improves the basic understanding of disease spread and results. Most of the cases are mild or asymptomatic, and asymptomatic cases do not show any symptoms. Still, after some time, presymptomatic cases show disease symptoms; all of these scenarios are considered in the proposed model. We have the number of total hospital beds of Mumbai city; based on that, we have calculated the number of beds required in the peak days of infection. Basic reproduction number and case fatality rate are the basic measures for this epidemic that we have also calculated. Sometimes conditions get worse because of a massive number of infected people at the same time, and the country does not have facilities to treat all critical cases when triage conditions occur. This model can solve the problem of triage. Depending on the current epidemic flow, there will be no need for new hospital beds, but if the next wave comes, this kind of scenario is not covered in this work.

## Limitation

Modeling is one of the most powerful tools that give intuitive effects when multiple factors act together. No model is perfect. Models are always simplifications of the real world. No models are perfect; there are some shortcomings in it. Proposed model also has some limitations, which are as follows: Our system of differential equations is very sensitive to initial parameters. We have to very careful while given the initial parameters. Small changes in parameters can cause a massive difference in results. Our present manuscript has especially focused on severe cases and death cases. We have consented the severe cases that do not get treatment in the critical compartment. We have considered recovered cases not infected again in the future. R0 value cannot be increased; it either decreases or remains constant. We have assumed that the cases that have been recovered will be immunized, meaning that they will not be infected again.

## 5 Summary

In this present manuscript, we have discussed the new model that is an extension of the SEIR-Model in which four major compartments are added that are Asymptomatic infectious, Quarantine hospitalized, presymptomatic infectious, and death compartment. Some people have not seen any symptoms of the disease. They are going to the asymptomatic infectious compartment. The proposed model estimates the date and magnitude of peaks of corresponding to exposed people, number of asymptomatic infectious people, presymptomatic infectious people, symptomatic infectious people, and the number of people hospitalized, the number of people quarantine hospitalized, and the number of death of COVID-19. A proposed model also calculates the basic reproduction number over time, and its computational results are almost the same as the actual data. According to the proposed model, Mumbai’s case fatality rate is very high, so the death cases are very high there, and with cases that are more critical.

For model validation, we have used three precision measures, MAPE, RMSE and R-squared. The proposed model outperforms the classic ARX model and the ARIMA model. In addition, it outperforms the deep learning model LSTM, and Seq2Seq model. To show the performance of the proposed model compared to the ARX and ARIMA models, SARIMAX, LSTM, and Seq2Seq model. To validate the performance of the mode Taylor diagram are included in the result section.

## Data Availability

Data is available online

https://covidindia.org/open-data/

## Conflicts of Interest

The authors have no conflicts of interest to report regarding the present study.

## Acknowledgement

I would like to express my special thanks of gratitude to our collaborator Harel Dahari of The Program for Experimental and Theoretical Modeling, Division of Hepatology, Department of Medicine, Stritch School of Medicine, Loyola University Medical Center, Maywood IL, United States and as well as Jonathan Ozik from Consortium for Advanced Science and Engineering, University of Chicago, Chicago, IL, USA, who gave me the golden opportunity to do work with them on SPARC project that helped us in doing a lot of good Research and we came to know about so many new things I am really thankful to them. We also thank Mrs. Ashwini Bhide, AMC, MCGM for her insights and for her crucial data inputs. We also thank Shri Saurabh Vijay, Secretary, Higher & Technical Education Department, and Government of Maharashtra for his insights and data inputs.

The work has been supported by a grant received from the Ministry of Education, Government of India under the Scheme for the Promotion of Academic and Research Collaboration (SPARC) (ID: SPARC/2019/1396).

## Notes

### Competing Interest Statement

The authors have declared no competing interest.

### Summary of Updates

Some references added

